# A Chemical and Digital Analysis of E-Cigarettes Seized from English Schools and the Social Media Marketplace

**DOI:** 10.1101/2025.09.04.25334826

**Authors:** Rachael Andrews, Ashley Fuller, Gyles E. Cozier, Matthew Gardner, Richard Bowman, Oliver B. Sutcliffe, Peter Sunderland, Tom P. Freeman, Gillian Taylor, Shane D Johnson, Fiona Spargo-Mabbs, Stephen M. Husbands, Jennifer Scott, Prianka Padmanathan, Christopher R. Pudney

## Abstract

**Background and Aims:** The use of synthetic cannabinoids (SCs) in e-cigarettes represents a significant public health threat to young people. Our previous research in the 2023-2024 academic year established a high prevalence of SCs in e-cigarettes seized from English schools and suggested that many of these were sourced through social media. This study provides a crucial update of drug occurrence in secondary schools for the 2024-2025 academic year and presents the first UK empirical evidence using digital content analysis of the availability of these substances on social media.

**Design and Methods:** E-cigarette and e-liquid samples were collected from a diverse cohort of 114 secondary schools across seven police force areas in England. Chemical analysis was performed using liquid chromatography-mass spectrometry (LC-MS) and quantitative nuclear magnetic resonance (qNMR) spectroscopy to identify and quantify controlled substances. This was supplemented by a survey of social media platforms to assess the online marketplace for these products.

**Findings:** Analysis of 1,933 samples revealed an average SC prevalence of 12.9% across the sampled regions, though this varied significantly between areas (1.2% to 27.0%). SCs were overwhelmingly found in refillable devices (21.1% positive) and unlabeled bottles (96.3% positive), with a very low prevalence in single-use products (0.9%). The study also identified the emergence of other controlled substances, including ketamine and MDMA. Digital content analysis confirmed a pervasive online market where SCs are deceptively marketed and sold as ‘THC’ vapes, particularly on platforms popular with younger demographics.

**Conclusions:** SCs remain a persistent threat in English schools, with consumption patterns strongly linked to refillable vape devices. Legislation targeting single-use vapes is unlikely to mitigate this issue. The deceptive mis-selling of SCs as THC on social media represents a major pathway to harm for young people. These findings underscore the urgent need for targeted harm reduction strategies in schools, greater accountability for social media platforms, and localized public health interventions to address this evolving challenge.

---

The use of e-cigarettes in by young people in Great Britain is rising, with 5% of 11-15 year olds and 12% of 16-17 year olds currently using e-cigarettes [1]. Concomitant with the rise in popularity of vaping is the availability of ‘cannabis’ (THC) e-cigarettes. These may be either a legal commercially available product (as local laws and regulations permit) or produced as an illegal product. Indeed, a third of people under 17 have used cannabis (2). The evolving social view of cannabis use is that it is an unremarkable feature of adolescent life (3).

In a recent study (4), exposure to e-cigarette or cannabis posts on social media was found to be associated with adolescent e-cigarette, cannabis, or dual use. The logical conclusion from this work is that the presence of these e-cigarettes on social media directly contributes to their use by young people. The vast majority of UK secondary-school-aged children use social media, with 91% of 12-15 year-olds and 98% of 16-17 year-olds having a social media profile (5). Therefore, the availability of THC e-cigarettes on these platforms would be an enhanced risk to young people. We note that in the UK THC is illegal and the advertisement for sale of THC (or any substance listed under the Misuse of Drugs Act 1971 or the Psychoactive Substance Act 2016) is a criminal offence, regardless of the identity of the advertised substance. The Online Safety Act 2023 makes explicit the responsibility of online platforms to protect young people from harm, including the advertisement and sale of illegal drugs.

Synthetic Cannabinoid Receptor Agonists (SCRAs; SCs), commonly known as ‘Spice’ or ‘K2’, represent a diverse and rapidly evolving class of New Psychoactive Substances (NPS) (6, 7). Despite being designed to mimic the effects of Δ9-tetrahydrocannabinol (THC), the primary psychoactive component in cannabis, SCs are pharmacologically distinct. They often act as highly potent, full agonists of cannabinoid receptors, in contrast to THC, which is a partial agonist (8, 9). This pharmacological difference contributes to a significantly higher risk profile (10). While acute cannabis toxicity is more commonly associated with cardiovascular symptoms like tachycardia and hypertension, SC use is linked to more severe neuropsychiatric effects, including drowsiness, coma, agitation, and seizures (11, 12). Indeed, case reports of adolescents using SC-laced vapes specifically document severe agitation and psychotic features (12), and the overall risk of requiring emergency medical treatment is estimated to be 30-fold greater for SCs compared to cannabis (10).

Historically in the UK, SCRA use was considered to be concentrated within specific vulnerable populations, such as people in prison and those experiencing homelessness (13, 14). However, national media and public health reports increasingly highlight a disturbing trend of SCRA-related health incidents among school-aged children, often linked to vaping (15). Our previous work provided the first systematic chemical analysis of this issue, revealing an alarming prevalence of SCs in e-cigarettes seized from English schools during the 2023-2024 academic year (16). We found that SCs were present in over 17% of all tested samples and were identified in more than three-quarters of the schools that submitted samples (16). This led us to hypothesise that young people, often intending to purchase THC vapes, were being mis-sold cheaper (17), more dangerous SC-laced products instead.

Given our initial findings and the evolving policy and online retail landscape, a follow-up study where new geographic regions are sampled is critical to growing the understanding of this issue. Therefore, this study presents a new analysis of e-cigarettes seized from a cohort of English secondary schools during the 2024-2025 academic year, as well as companion digital content analysis of the landscape of online availability of the material via social media. We aim to identify and quantify the psychoactive substances present, establish strategies for identifying SC containing material, ultimately to provide evidence to inform harm reduction strategies, public health policy, and enforcement priorities.

## Materials and Methods

### Selection of Geographic Regions

The study focused on geographic areas where the regional police force or council were concerned about incidents of synthetic cannabinoid (SC) intoxication or where an assessment of occurrence was desirable from a public health perspective. We secured the necessary permissions for sample collection and delivery to our laboratory. Formal agreements were established for four distinct areas. These include testing with four police force areas, Devon and Cornwall, Essex, Lancashire and Merseyside. We report only the broad geographic area of the sampling effort to provide anonymity to precise testing locations (e.g. cities) at the request of the police forces or local councils involved in the study. For other regions where requests were made, either the relevant authorities chose not to participate within the study’s timeframe or permission was not granted.

### Sample Collection and Handling

The local police force invited schools in their jurisdiction to voluntarily submit e-cigarette and e-liquid samples that had been confiscated by staff. The submission process was designed to be unbiased; no specific schools were targeted, and teachers followed their standard procedures for confiscating items. All samples were collected during the 2024/2025 academic year. The police forces served as the central point for collating all submissions from the schools without any secondary selection. Samples were transported by the police to our laboratories for detailed analysis. Each submission was queried for sample bias, for example clarifying if samples were submitted associated with known health incidents and if so were removed from the data set presented here.

All samples were visually inspected to document their form factor; Single-Use (SU), Refillable (RF), Labelled Bottle (LB), or Unlabelled Bottle (UB), and the color of the e-liquid.

### Analytical Procedures

Samples were analyzed using liquid chromatography–mass spectrometry (LC-MS) to screen for substances controlled under the Misuse of Drugs Act (1971) and the Psychoactive Substances Act (2016). Identification was performed by matching against the HighResNPS and ForTox mass spectral libraries as we reported previously (16).

For LC-QToF-MS analysis, all e-cigarette liquid samples were initially diluted 20 000x in HPLC grade ethanol, with repeats at higher concentration where needed/possible. Analyses were performed using an Agilent QToF 6545 with a Jetstream electrospray ionization (ESI) source coupled to an Agilent 1260 Infinity II Quaternary pump HPLC with a 1260 autosampler, column oven compartment, and variable wavelength detector (VWD). The mobile phases used were (A) LC-MS grade water with 0.1% formic acid and (B) acetonitrile with 0.1% formic acid. The gradient used was 95:5 A:B from 0.00-0.60 min, change to 0:100 A:B over 0.60-3.00 min, held at 0:100 A:B from 3.00-5.50 min, change to 95:5 A:B over 5.50-5.60 min, and held at 95:5 A:B from 5.6-7.6 min. The flow rate was 0.5 mL/min at 50ºC and 5 μL of the sample was injected onto an EC-C18 3.0 x 50 mm, 2.7 μm particle size column (InfinityLab Poroshell 120, Agilent Technologies). The MS was operated in positive ionization mode with the gas temperature at 250°C, the drying gas at 11 L/min, and the nebulizer gas at 35 psi (2.41 bar). The sheath gas temperature was set to 300ºC and the flow rate was 12 L/min. The MS was calibrated using a reference calibrant introduced from an independent ESI reference sprayer. The VCap, Fragmentor and Skimmer were set to 3500, 160, and 45 V, respectively. The MS was operated in all-ions mode with three collision energy scan segments at 0, 20, and 40 eV.

The VWD was set to detect at 280 nm wavelength at a frequency of 2.5 Hz. Data processing was automated in Qual 10, with the molecular feature extraction set to the largest 20 compounds for [M^+^H]^+^, [M-H]^-^, and [M+HCOO]^-^ ions. The results were also searched against the online mass spectral databases HighResNPS (containing over 2300 unique compound entries) and ForTox, with a forward score of 25 and reverse score of 70, and mass tolerances within 5 ppm of the reference library matches. Qualified ions had co-elution scores of ≥ 90, retention time tolerances of ± 0.10, and a minimum S/N of ≥ 5.00.

To quantify the amount of SC present, quantitative NMR (qNMR) was used. The method used was based on a study that quantified SCs in seized e-liquids.^18^ Samples were prepared by mixing 200 μL of e-cigarette liquid with 400 μL of methanol-d4 (MeOD) containing 3 mg of 3-(trimethylsilyl)propionic-2,2,3,3-d4 acid sodium salt (purity ≥ 99%, isotopic purity 98 atom % D) (TSP). A set of MDMB-4en-PINACA concentrations (10 – 0.1 mg/ml) were prepared in 50/50 propylene glycol/glycerol (PG/VG) as standard e-cigarette liquids to test the quantification method.

^1^H NMR data were recorded on a Bruker AvanceCore 400 MHz spectrometer (1H frequency of 400.130), with a zg pulse sequence composed of 3.18 s acquisition time, 128 scans and 20 s delay. Chemical shifts were referenced to 3.31 ppm for residual CD_2_HOD solvent peak (from MeOD) and are reported in ppm. NMR spectra were processed with Mestralab Mnova 14.1 using automatic phase and Whittaker smoother baseline corrections, followed by zero filling (4 x original size) and line broadening (1 Hz) to improve signal/noise ratio. Due to the large amounts of PG/VG in e-cigarette liquid, only the 4 peaks from the aromatic indazole core of the SCs could be reliably integrated. As many of these were used for the qNMR calculation as possible, when not obscured by other additives in the e-cigarette liquid.

The following equation was used for the ^1^H qNMR quantitation:

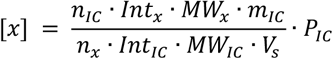

Where [*x*] denotes concentration in mg/mL, P is the purity, n is the number of protons, Int is the integral value, MW is the molecular weight, m is the mass in mg, V is the volume in mL, IC is the internal calibrant, x is the analyte, and s is the sample. As the indazole peaks of the different SC compounds over-layed, when multiple SC compounds were present in each sample, the molecular weight of the main SC, as judged by the LC-MS chromatogram, was used in the calculation.

We have sought permission to report the location of the sampling regions as the broad police force area where the sampling took place. That is, the data do not reflect sampling across the entirety of each region described, respecting that specific cities or regions may not want negative attention associated with reporting of the data.

### Digital content analysis

We selected three major social media platforms (*Facebook, TikTok* and *Instagram*) where specific user accounts and their details can be searched for and viewed, including photos and videos. Digital content analysis was performed on the social media platforms on 18th June 2025 and then every month for two months (ending August) for TikTok and Instagram. Facebook was not monitored over this period based on specific platform limitations as described in the main text below (Results and Disucssion). Accounts were manually identified using the search terms ‘THC Liquid’, ‘THC Liquid UK’, ‘spice liquid’, ‘spice liquid UK’, ‘K2 liquid’, and ‘K2 liquid UK’.

It is illegal for academic researchers in the UK to test purchase illicit drugs from the internet and so our visual triage approach is the only possible route. As we discuss in the main text (Results and Discussion), our findings allow us to establish accurate visual criteria for classification: SCs were identified by (i) unlabelled bottles of liquid, (ii) video evidence of a low-viscosity liquid (not an oil/resin), or (iii) visual evidence of solid SC powder associated with manufacture. THC was identified by (i) commercial packaging associated with known THC e-cigarette brands or (ii) visual evidence of a viscous yellow/brown oil. Based on recent analysis (17), price was also used as a contextual factor, with liquids priced at ~£2-10 / mL considered likely SCs and those >£10 / mL considered likely THC. Accounts were identified as showing SCs even if THC was additionally verified by the visual triage. Based on the visual triage of the advertised products, each account was classified by the research team as likely selling synthetic cannabinoids (SCs), THC, or as ‘unclear’ – including showing no image/video file to assess.

The bio or description text for each classified account was then extracted for linguistic analysis. We have removed any text from the reported accounts that would lead to identification of the accounts. An inductive thematic analysis was employed to identify key marketing themes. This involved an initial familiarization phase where all text was read multiple times, followed by a systematic coding process to label recurring keywords and phrases. These codes were then collated and organized into broader themes which were reviewed and refined. The specific themes are described in the main text. To determine if the observed differences in language use between the SC and THC groups were statistically meaningful and between social media platforms, Fisher’s Exact Test was employed, which is appropriate for comparing categorical variables between groups with small or uneven sample sizes.

### Reflexivity statement

We were conscious that our familiarity with drug market terminology and our previous work could influence the coding process. To mitigate this, we adhered strictly to an inductive thematic analysis, allowing themes to emerge directly from the sellers’ own language before applying our external knowledge. As such peer debriefing was conducted to review the codes and themes.

## Results and Discussion

Table 1 gives the data for all of our sampling efforts to date, including those reported by us previously from the 2023/2024 academic year (16). Figure 1A shows the SC percentage data (Table 1) mapped for each sampled region. We have also included representations for areas where we have identified SC e-cigarettes or e-liquids submitted from a secondary school in that region (black shaded areas in Figure 1), but where we have not performed volume sampling. These data show a more regionally diverse sampling picture compared to our sampling from the academic year 2023/2024. Together, the average percentage of SC material is 9.5 %. However, different regions are sampled to different depths – for example Essex has 746 samples, versus 84 from greater Manchester. It is therefore more appropriate to calculate the average occurrence from the regions, giving an average of 12.9 ± 9.5 % (Table 1). The large standard deviation demonstrates the variability in the occurrence of SCs. Comparing the occurrence of SCs to THC illustrates a stark difference. With the average percentage of samples being THC across the tested regions being 1.2 ± 1.1 % (Table S1). That is, ten times lower than the occurrence of SC samples. Images of all positive samples are given in Figure S1 and key illustrative examples shown in Figure 2.

**Table 1.** Summary of analysis of e-cigarettes and e-liquids seized from English secondary schools.

|  | Total samples<br>(no. Schools) | SU – Single use | RF - Refillable | LB – Labelled<br>e-liquid | UB – Unla-<br>belled e-liquid | SC present |
| --- | --- | --- | --- | --- | --- | --- |
| London (23/24)<sup>a</sup> | 156 (13) | 106 – 67.9% | 27 – 17.3% | 11 – 7.1% | 12 – 7.7% | 36 – **23.1%** |
| South Yorkshire<br>(23/24)<sup>a</sup> | 270 (9) | 200 – 74.1% | 34 – 12.6% | 20 – 7.4 % | 19 – 7.0% | 40 – **14.8%** |
| Greater Manchester<br>(23/24)<sup>a</sup> | 84 (6) | 37 – 44.0% | 28 – 33.3% | 10 – 11.9% | 9 – 10.7% | 14 – **16.7%** |
| Merseyside (24/25) | 207 (14) | 176 – 85.0% | 21 – 10.1% | 4 – 1.9% | 0 – 0% | 2 – **1.2%** |
| Devon and Corn-<br>wall (24/25) | 255 (34) | 150 – 58.8% | 62 – 24.3% | 37 – 14.5% | 6 – 2.4% | 14 – **5.5%** |
| Essex (24/25) | 746 (30) | 554 – 75.3% | 125 – 17.0% | 45 – 6.1% | 11 – 1.5% | 19 – **2.5%** |
| Lancashire (24/25) | 215 (8) | 62 – 28.8% | 97 – 45.1% | 31 – 14.4% | 25 – 11.6% | 58 – **27.0%** |
| Total - % of total | 1933 (114) | 1285 – 66.5% | 394 – 20.4% | 158 – 8.2% | 82 – 4.2% | 183 – 9.5% |
| Region average<br>(%)<sup>b</sup> | - | 61.9 ± 18.1% | 22.8 ± 11.6% | 9.0 ± 4.4% | 5.8 ± 4.3% | **12.9 ± 9.5%** |

a, Data reported in Cozier *et al.* 2025 (16). b, data reported as mean ± standard deviation.

**Figure 1.**
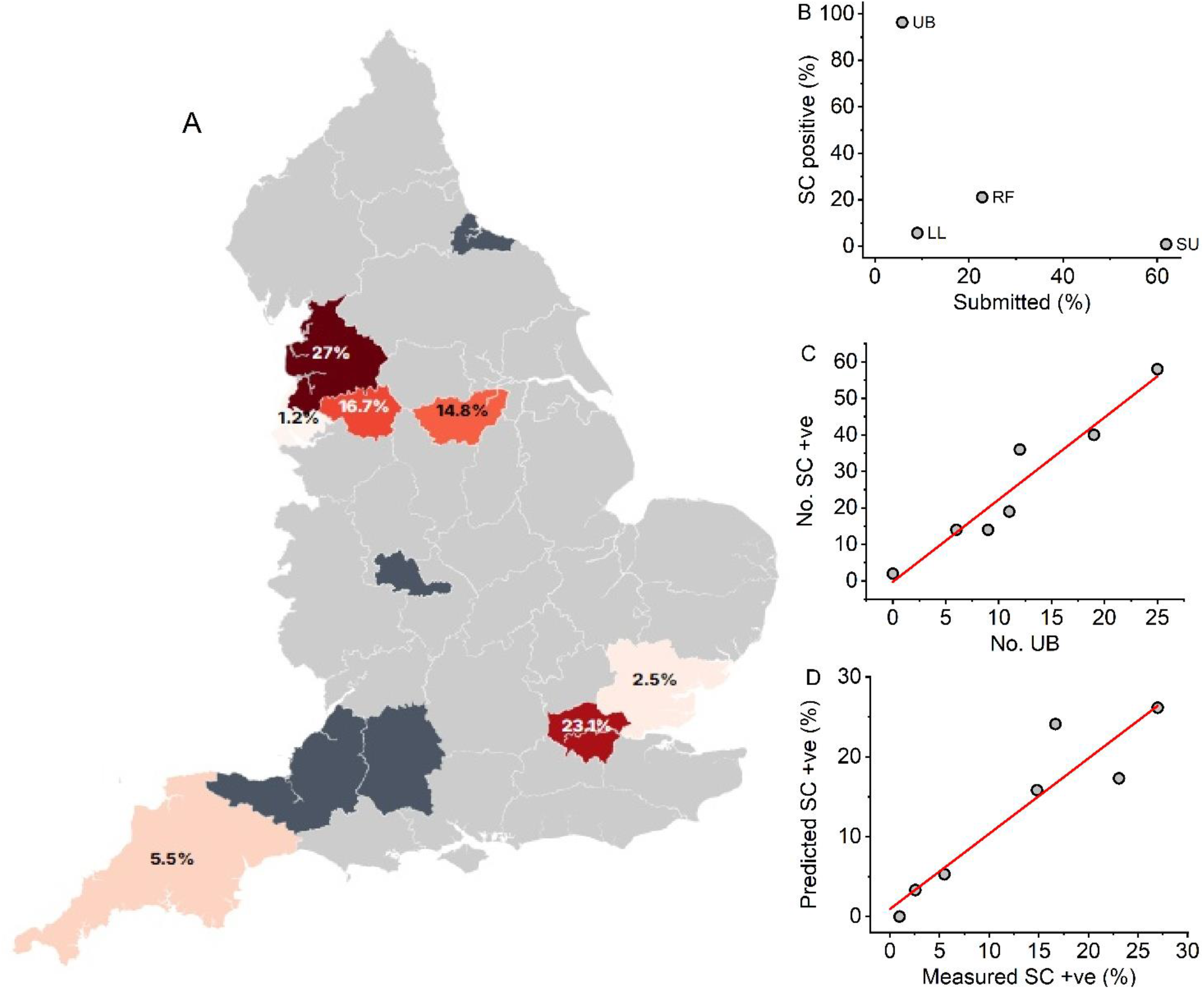
Occurrence of SC e-cigarettes and e-liquids in England. **A**, A geographic map representing measurement and sampling efforts of e-cigarettes and e-liquids from secondary school in England. Colored regions represent sampling efforts given in Table 1. Black regions represent areas where we have identified SC material seized from secondary schools, but have not completed an at scale sampling effort. Percentages indicate the proportion of schools with a positive test for an SC. **B**, The percentage of physical sample types with SCs present *versus* their percentage of submission. **C**, Correlation between the number of SC positive samples and the number of UBs. **D**, “Validation of the prediction model: Correlation between the predicted percentage of SC positive samples (calculated from the number of UBs and the analytically measured percentage.

**Figure 2.**
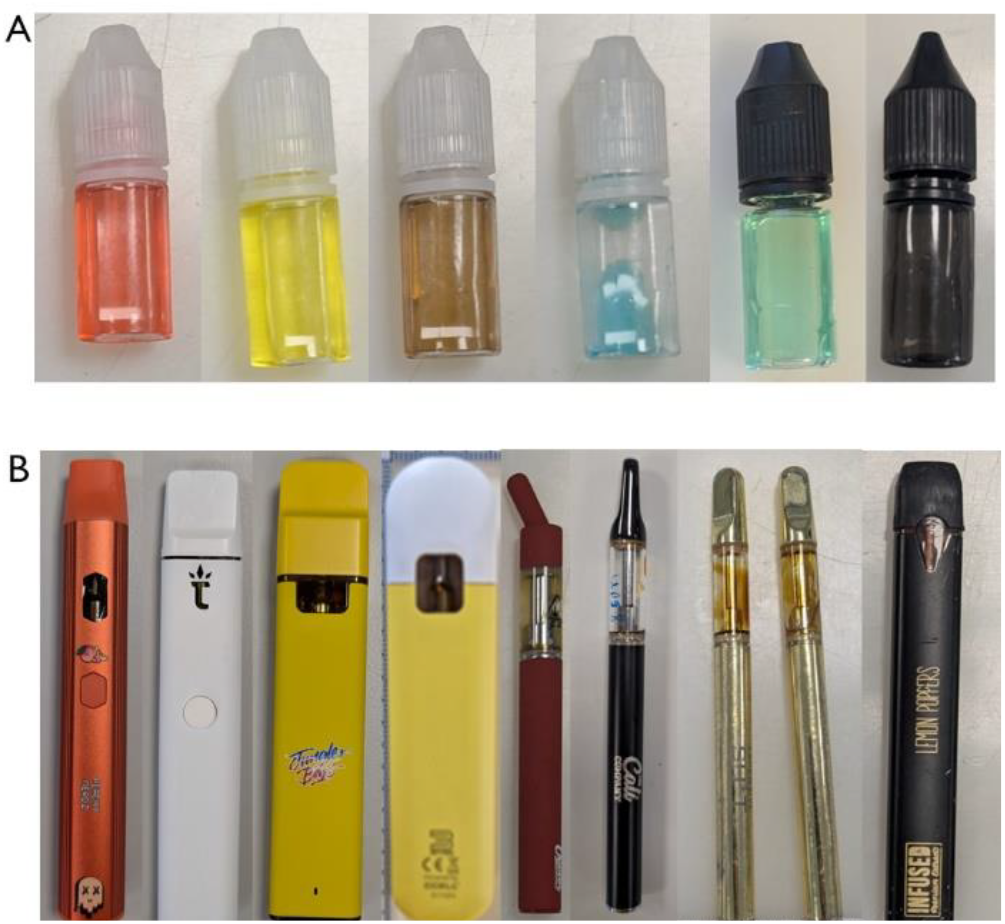
**A and B**, Examples of SC e-liquids (panel A) and THC e-cigarettes (panel B).

We note that we have sampled Devon and Cornwall and Essex twice throughout the 2024/2025 academic year and the data are the sum of those sampling efforts. The data from each exercise are given in *Supporting Information* (SI) Table S1. From Table S1, the data are similar between sampling efforts (approximately 6 months apart) for these regions, with the percentage of SC positive samples being 7.8 % and 4.9 % for Devon and Cornwall and 2.9 % and 2.4 % for Essex. That these temporally different sampling efforts produced consistent percentages of SC positives, suggests that the overall landscape of SC use (at least for these regions) is not highly variable within an annual cycle.

Liverpool (a city within the Merseyside police area), is a major national hub of county lines drug trafficking (18). We have therefore queried if the very low percentage of SC finds in Merseyside (1.2%) is corroborated by other policing intelligence. To that end we asked if Merseyside Police were aware of any incident’s involving vapes in schools within the last two years. The Intelligence, News and Communications teams in this force area had no alerts associated with e-cigarettes in this time frame. That is, despite Merseyside having a high prevalence of drug dealing and abuse compared to the national average (19), they do not have a high occurrence of SC e-cigarettes or liquids being found within secondary schools. This is juxtaposed with the same reporting from Lancashire Constabulary where SC finds were high (27%). They report over 181 youth related vaping incidents between January 2024 and October 2024 in the specific testing region of the force area. Moreover, the number of these incidents approximately doubled over the time period.

Our data do not inform why geographically close regions can have significantly different occurrences of SC e-cigarettes. Our previous data have suggested a weak positive trend with a single metric of social deprivation (the fraction of free school meals; FSM). Figure S2, shows this same trend now with the further data from sampling, plotting the data for schools where >20 samples were submitted. From a Pearson’s correlation analysis, P = 0.097 and r = 0.444. While a moderate positive trend is observed, it does not reach statistical significance, suggesting that social deprivation, as measured by FSM, is not the sole explanatory factor for the regional variance

Figure 1B shows the percent of submitted e-cigarette and e-liquid types *versus* the percentage of those samples containing SCs. These data show (and given in Table 1) that by far SU e-cigarettes are the most submitted (61.9 ± 18.1%) and UBs the least common, 5.8 ± 4.3%. However, 96.3 % of UBs and 21.1 % of RFs contained SCs. Only 0.9 % of SUs and 5.7 % of LBs contained SCs. These data mirror our previous findings (16) that SCs are overwhelmingly used in RFs, most likely filled using UBs. The median and maximum concentration of the samples from 2024/2025 was 0.65 and 3.7 mg / mL, respectively. This is essentially the same as our 2023/2024 findings; 0.42 and 3.6 mg / mL. Given the concentration measurements from the two sampling exercises are essentially the same argues for some consistency of manufacture of these samples or potentially a limit of solubility of SCs in e-liquids.

Given the extremely high occurrence of SCs, and the conceptual link to SCs in UBs, we were interested to explore if the simple visual identification of UBs could be used as a predictor for SC occurrence in a sample set. Figure 1C, shows the number of SC positives *vs* the number of UBs from each region (Table 1). The solid red line is the fit to a simple linear equation, which suggest a positive correlation, having a gradient of 2.25 ± 0.29 and a y intercept at −0.2 ± 4.0. These data are statistically significantly correlated with a Pearson’s correlation coefficient, r = 0.962 and p = 0.0005. That is, the number of SC positives from all samples, is accurately predicted from the number of UBs, simply calculated by multiplying the number of visually identified UBs by the gradient of the fitted line in Figure 1C (2.25). Figure 1C shows the correlation between the predicted percentage of SC positives using this method and the actual measured percentage of positives using our analytical approaches (described above). The solid fitted line again shows a positive correlation, with a Pearson’s correlation coefficient, r = 0.928 and p = 0.0026. These data demonstrate that with sufficiently deep sampling of a region, the visually identified number of UBs can be used as an accurate proxy for the occurrence of SCs in e-cigarettes and e-liquids, at least when the material is from an English secondary school.

Part of our motivation for further sampling of e-cigarettes and e-liquids from schools was to monitor for any evolution in the landscape of illicit drugs being used in secondary schools. For example, in our previous sampling effort, we found a single sample with heroin present. From the new testing reported here we have identified two samples containing ketamine and one containing MDMA. One ketamine and MDMA sample were identified in Devon and Cornwall on the first sampling effort for this region (Table S1 and Figure S1). The second ketamine sample was identified in Lancashire (Table S1 and Figure S1). We have quantified these samples and we find that they are present at less than 0.1 mg / mL. That is, they represent a very low dose compared to the typical expected use of these drugs.

### Digital content analysis of SC availability on social media

The findings from our sampling study (Table 1; Figure S1) are highly consistent in that THC e-cigarettes present in a different form factor to SC e-cigarettes and liquids. That is, to a reasonable level of approximation, one can visually distinguish between THC and SC e-cigarettes and e-liquids and we describe the criteria in detail in *Materials and Methods*. Briefly, THC e-cigarettes are typically in a ‘slim’ e-cigarette designed to hold a small volume of material (~1 mL) and visually show a yellow/brown oil/resin via a viewing port to the heating coil. In contrast, SC e-liquid is almost always in an unlabeled bottle (~10 mL) and can be a broad range of colors. We show this generic distinction in Figure 2. Indeed, our data (Figure 1B) show that 96.3 % of UBs are an SC (never THC) and that visual identification of UBs can accurately represent the fraction of SC from a surveying effort (above).

Given these data, and the apparent ability to visually triage THC *vs* SC material as it presents in material seized from schools, we were motivated to survey the availability of these on selected social media platforms. We applied simple keyword searches as described in *Materials and Methods*. We then identified and logged the accounts where there was evidence that either SCs, or THC was putatively available. These data are given in Table 2 and example images from the accounts are shown in Figure 3.

**Table 2.** Identification of accounts via key word searches on three social media platforms.

|  | Facebook | Instagram | TikTok |
| --- | --- | --- | --- |
| "THC liquid (UK)" |  |  |  |
| Total | 75 | 83 | 120 |
| THC - (%) | 46 – 61.3% | 6 – 7.3% | 20 – 16.6% |
| SC - (%) | 9 - **12.0%** | 45 – **54.8%** | 81 – **67.5%** |
| Unclear - (%) | 20 – 26.7% | 32 – 39.0% | 19 – 15.8% |
| "Spice/K2 liquid (UK)" |  |  |  |
| Total | 50 | 20 | 18 |
| THC - (%) | 0 | 0 | 0 |
| SC - (%) | 50 – 100% | 20 – 100% | 11 - 61.1% |
| Unclear - (%) | 0 | 0 | 7 - 38.9% |

**Figure 3.**
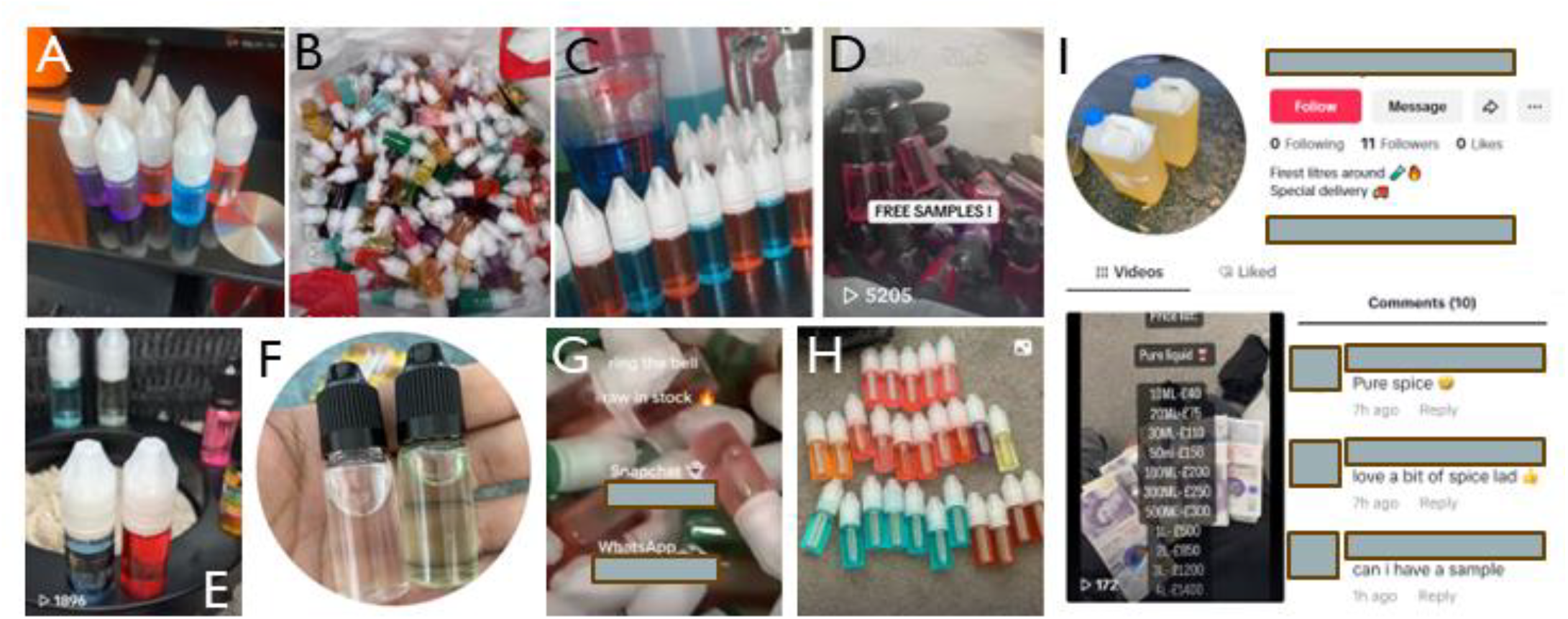
Examples of samples used to putatively identify SC liquids. Panels **A-I** represent different accounts from either TikTok or Instagram. Panel **I** shows further details from a specific account including the acknowledgement the products are SCs from comments and pricing structure.

From Table 2, we find that when directly searching for SC material (‘spice/K2 liquid (UK)’), we identify 88 accounts across all three platforms offering to sell SC material in some form. We note that these accounts are almost always combined with the sale of SC-soaked sheets of paper intended for a prison environment (20). In this case the ‘liquid’ is intended to soak the sheets of paper and we do not suggest they represent liquid suitable for an e-cigarette, nor is it marketed for that purpose. We note that for Facebook, the number of accounts purporting to sell spice/K2 was large (> 1000; including people and groups) and so we identified the first 50 in the search list for parity with the sampling of other platforms.

From Table 2, using the search term “THC liquid (UK)” we find that different social media platforms give different ratios of putative THC and SC containing samples, with TikTok having the highest fraction (67.5 %) of putative SC material and Facebook the least (12 %). These data include the accounts where a determination of THC/SC could not be made. Similar to our searches for spice/K2 above, the number of accounts purporting to sell THC on Facebook was high (>1000 including people and groups for “THC Liquid UK”) and so we identified the first 75 in the search list for parity with the sampling of other platforms. For TikTok and Instagram, the search term “THC liquid (UK)’ returned examples that were focused almost exclusively on e-cigarettes and e-liquids, with only occasional edible products. Given the accounts identified from Instagram and TikTok are directly relevant to this study and the Facebook accounts are less relevant, our subsequent content analysis focusses on these two platforms. Moreover, the account descriptions on Facebook were not sufficiently detailed to allow us to complete the thematic analysis below.

Tables S2 and S3 show the text associated with each account from TikTok and Instagram that arose from the search term(s) “THC Liquid (UK)”. We have anonymized the account names and identifying information such as link addresses. From inspection of the text, 68.4% of TikTok and Instagram accounts identify as selling to the UK or offer world-wide/national shipping. Therefore, the data presented and analyzed here are relevant to the studies geographic sampling area. 11.9 % of the putative SC selling accounts offer a postal service. 46.3 % of accounts link to another source, with 32.0 % linking to the encrypted messaging platform Telegram and more rarely (8.9 %) to Snapchat. Remaining links were to WhatsApp (4), Signal (3) or Discord (2). Finally, seven of the SC positive accounts quoted a pricing or pricing structure (e.g. Figure 3I), with a median price of £30 / 10 mL, which is similar to a recent report monitoring webshops (17). No accounts apparently selling THC alone gave pricing information.

From the accounts, we identified videos showing apparent production of SC e-liquid, showing as the mixing of an apparent e-liquid in a ‘home lab’. We cannot confirm this represents production, but is indicative of potential local production (of the e-liquid) in the UK. We did not find any video or photographic evidence showing the organic synthesis of the SC.

A thematic analysis of the linguistic patterns in the descriptions of 203 social media accounts (83 from Instagram, 120 from TikTok) reveals a clear operational model for the online sale of illicit e-cigarettes. The thematic analysis coding framework is given in Table S4 and the categorization of the accounts shown in Tables S2 and S3. The two most dominant themes across the dataset were Funnelling to Private Channels (present in 52% of accounts) and a focus on the UK Market (68.4%). This shows a core strategy of using public-facing social media to advertise to a specific audience before moving them to less-moderated, encrypted platforms like Telegram for the actual transaction.

However, a comparative analysis reveals statistically significant differences in marketing strategy between the platforms (Table S4). Sellers on Instagram were significantly more likely to use language related to Product Form/Authenticity (p<0.0001; 30.1% of Instagram and 5.0 % of TikTok accounts), using terms like ‘flavours’, ‘Cali’, or ‘resin’, which we suggest is to build a more sophisticated brand image. In contrast, sellers on TikTok employed a more direct approach with significantly less emphasis on product branding. TikTok accounts were more likely to use Funnelling to Private Channels than Instagram (P<0.0496; 59.2 % of TikTok and 44.6% of Instagram accounts). While other themes such as claims of Extreme Potency or use of Plausible Deniability were present, their use was not statistically different between the two platforms. Similarly, we do not find any statistically significant differences in the themes between accounts identified as SC and THC.

Finally, we wished to assess the longevity of the tracked accounts. To that end we surveyed all tracked accounts on Instagram and Facebook on 01 September 2025 (the immediate end of data collection). We found that 74.7 % of Instagram and 68.3% of TikTok accounts were still active on this date. We did not find a clear link between the account name, description or content that might make these accounts targeted specifically for removal.

We acknowledge that the thematic analysis is based on a visual triage, not chemical analysis, is time bound over a relatively short period and limited to specific platforms. In summary, the analysis confirms that while sellers share operational similarities, their core marketing strategies show some difference. This suggests that different products are being targeted at different user demographics with distinct expectations.

## Conclusions

This study confirms that synthetic cannabinoids (SCs) remain a significant and persistent threat within English secondary schools and on social media in the academic year 2024-2025. Our analysis of seized e-cigarettes reveals a complex and regionally varied landscape of SC prevalence, with an average occurrence of 12.9% across the sampled regions. This represents a slight decrease from the previous academic year, which we suggest reflects the expanded sampling to more regions, or that the occurrence of this material has shifted in a regional manner during the timescales of the sampling efforts. However, the continued presence of these potent substances, identified in every surveyed region, alongside the emergence of other drugs like ketamine and MDMA in e-cigarette form, underscores an evolving and dangerous market targeting young people.

Our findings strongly indicate that the mode of consumption is a critical factor. SCs are overwhelmingly found in refillable devices and unlabeled e-liquid bottles, with single-use e-cigarettes rarely containing these substances. This suggests that legislation in England and Wales that has banned singleuse products is unlikely to impact the prevalence of SCs in schools and may inadvertently push users towards refillable options that are more likely to be use with illicit drugs. We have also established that the number of unlabeled bottles within a sample set can serve as a reliable proxy for the overall prevalence of SCs, providing a valuable tool for future monitoring.

While users of all ages can be found on each social media platform that we monitored, the core demographics for each are distinct. Facebook serves a broader, older audience, Instagram is typically used by young adults, and TikTok is the primary platform for the youngest generation of social media users. For example, in 2024 > 60% of US adolescents age 13-17 report ever us of TikTok or Instagram versus 32 % for Facebook (21). There is therefore an apparent relationship between the demographic that typically access each social media platform and the percentage of SC samples, sold as THC: From Table 2, Facebook (12%) – Instagram (54.2%) – TikTok (67.5%). That is, younger audiences are more likely to encounter SC material, sold as THC.

Our digital content analysis reveals a pervasive and deceptive online marketplace, which disproportionately targets younger people with counterfeit THC products that are, in fact, putatively SCs. We suggest that this mis-selling exploits the perception of cannabis as relatively safe and preys on the inability of young consumers to distinguish between the products, creating a direct pathway to harm. These findings are particularly timely as our social media data collection period - from June to August 2025 - spans the July 25th 2025 enforcement date for key provisions of the Online Safety Act 2023 (UK). This legislation makes explicit the responsibility of online platforms to protect young people from harm, including the advertisement and sale of illegal drugs. The fact that our simple keyword searches readily identified numerous accounts selling these substances during this period, and their apparent longevity, highlights a clear failure in existing content moderation. It provides a critical baseline showing that platforms are not proactively meeting their duties, underscoring the urgent need for robust and immediate enforcement of the new legislation to hold these companies accountable.

We point out that we have used extremely simple and limited search terms to identify the accounts presented in this manuscript. More complex and evolving terminology is likely to develop if these simple keywords are restricted and so we argue for a combination of text and graphical analysis to identify and restrict access to these accounts. This seems especially plausible given the high accuracy of our simple visual triage process and the evolving capabilities of artificial intelligence and machine learning models to discriminate images.

The implications of these findings are immediate and far-reaching. There is an urgent need for targeted, evidence-based harm reduction strategies within schools that focus on the specific dangers of refillable devices and unlabeled liquids. Public health campaigns must address the deceptive marketing of SCs as THC, and social media companies must be held accountable for the blatant sale of these illegal and harmful products on their platforms addressing clear failings in their content moderation systems and duties under current platform policies and national legislation. Finally, the regional disparities in SC prevalence highlight the need for continuous monitoring, localized policing and public health interventions to address this ongoing threat to the health and safety of young people.

## Supporting information

Supporting information to the main manuscript

## Data Availability

All data produced in the present work are contained in the manuscript.

## ASSOCIATED CONTENT

### Supporting Information

Example images of e-cigarettes and e-liquids. Data relating to testing. Data relating to social media searches.

## AUTHOR INFORMATION

### Author Contributions

The manuscript was written through contributions of all authors. All authors have given approval to the final version of the manuscript.

### Funding Sources

EPSRC (EP/V026917/1 and EP/L016354/1).

## ACKNOWLEDGMENT

We are grateful to the participating police forces and councils that enabled the work to be completed.

## Notes

### Competing Interest Statement

The authors have declared no competing interest.

### Funding Statement

This study was funded by EPSRC (EP/V026917/1 and EP/L016354/1).

