## Supporting information to the main manuscript for "A Chemical and Digital Analysis of E-Cigarettes Seized from English Schools and the Social Media Marketplace"

^1^Department of Life Sciences, University of Bath, Bath, BA2 7AY, UK. ^2^Dawes Centre for Future Crime, University College London, UK. ^3^School of Physics and Astronomy, University of Glasgow, Glasgow, G12 8QQ, UK. ^4^MANchester DRug Analysis & Knowledge Exchange (MANDRAKE), Department of Natural Sciences, Manchester Metropolitan University, Manchester, M1 5GD. ^5^Department of Psychology, University of Bath, Bath, BA2 7AY, UK. ^6^School of Health and Life Sciences, Teesside University, Middlesbrough, TS1 3BX, UK. ^7^Daniel Spargo-Mabbs Foundation, Palmerston House 814 Brighton Road Purley CR8 2BR, UK. ^8^Bristol Medical School, University of Bristol, Bristol, BS8 2PS, UK ^9^Centre for Academic Primary care, Bristol Medical School, University of Bristol, Bristol, BS8 2PS, UK. ^10^Centre for Bioengineering and Biomedical Technologies, University of Bath, Bath, BA2 7AY, UK.

KEYWORDS Synthetic cannabinoids, spice, K2, school, e-cigarette, vaping, THC

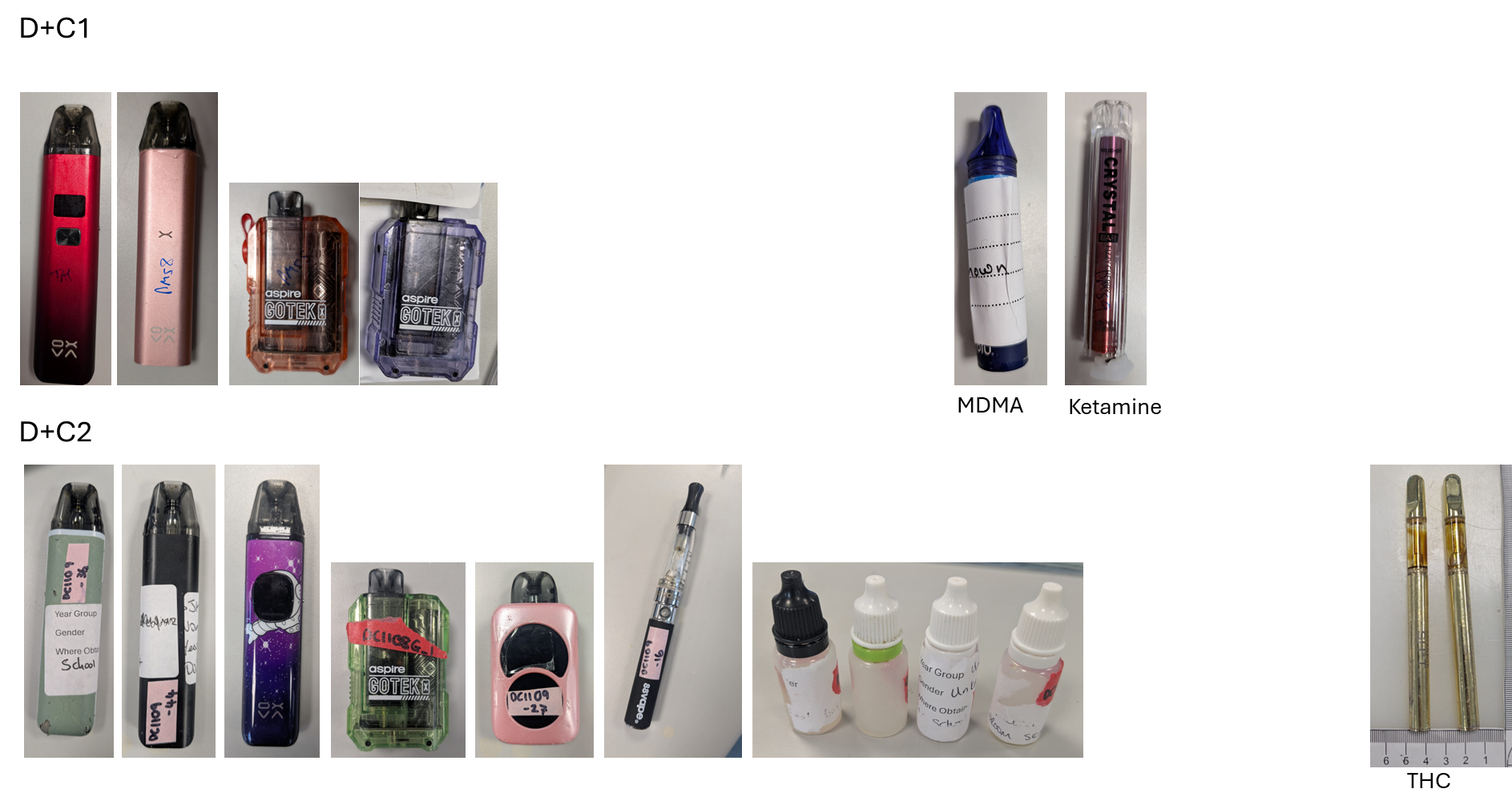

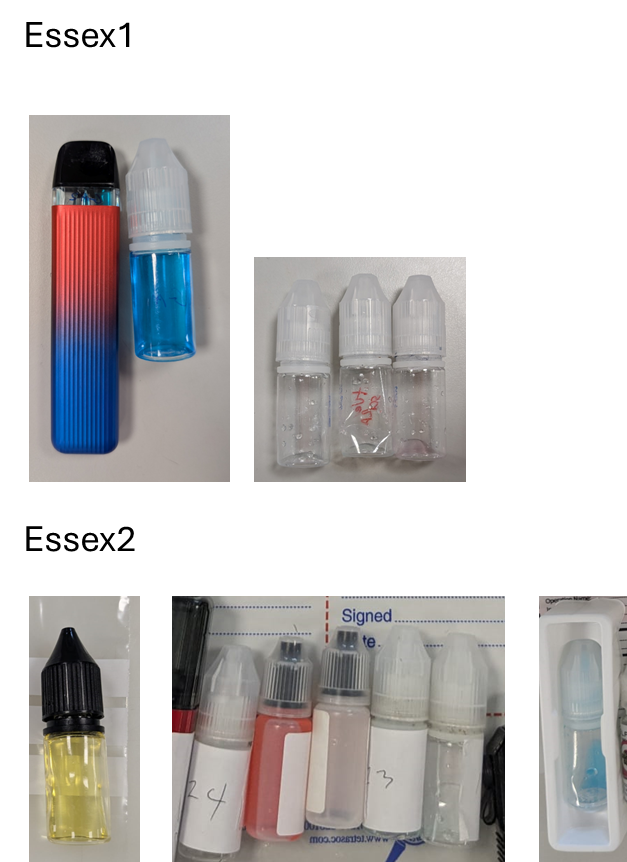

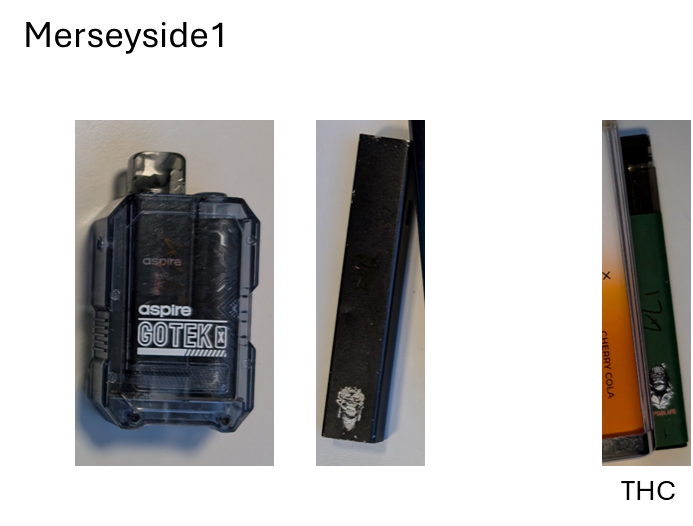

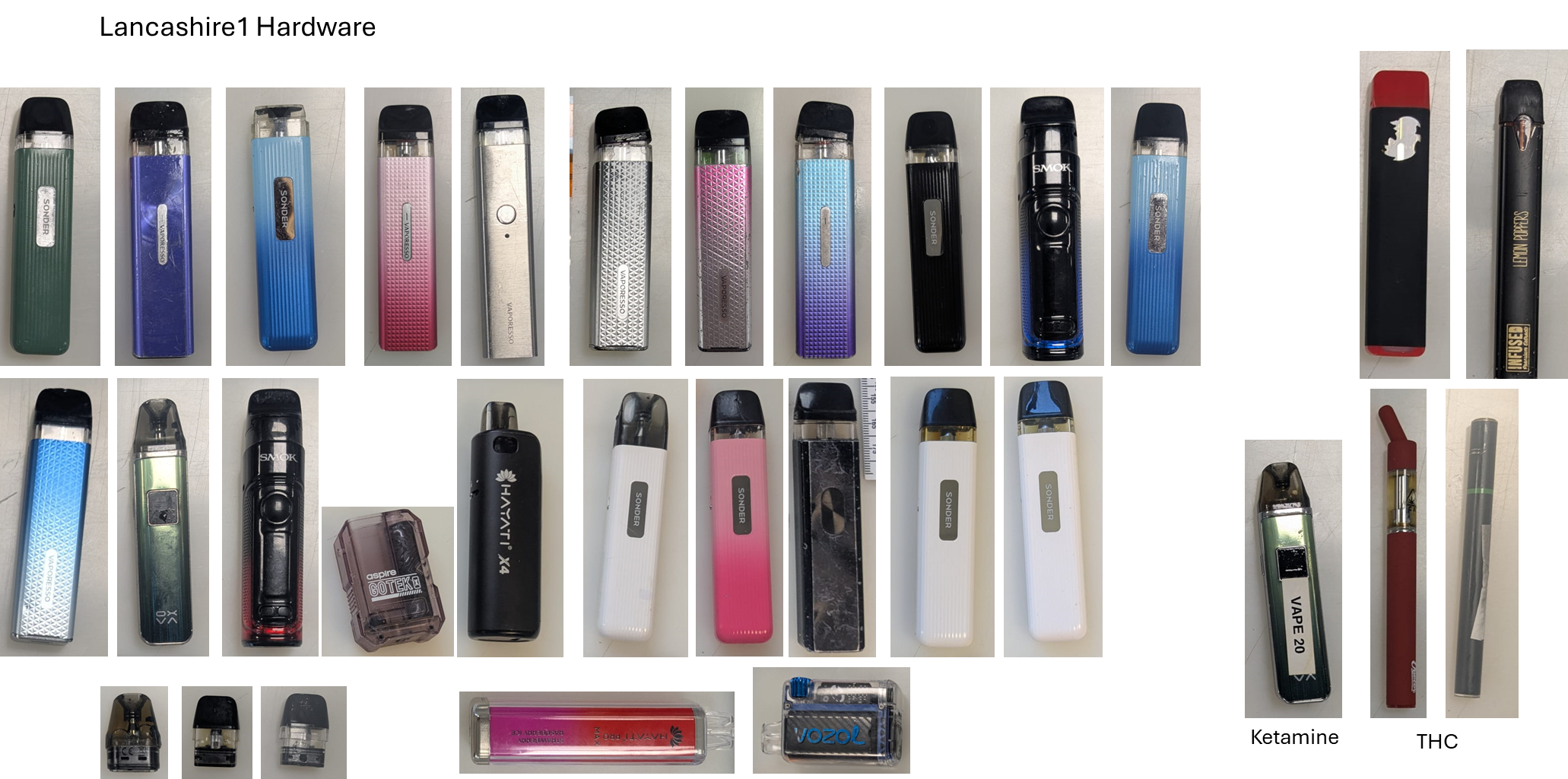

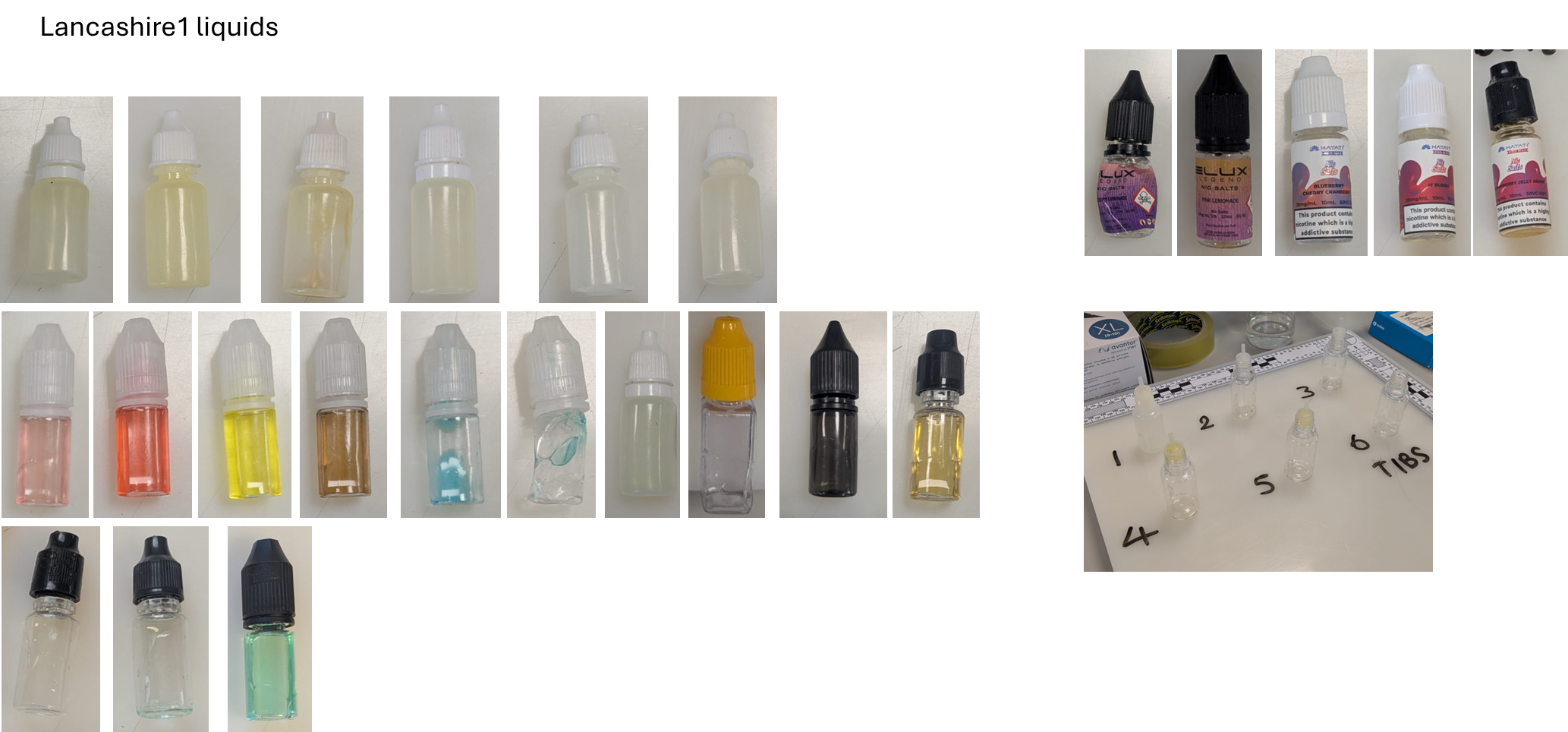

**Figure S1**. Summary of e-cigarettes and e-liquids found to contain an illicit drug as described in the main text. Samples are categorized as the heading above each image set. All samples contained SCs unless otherwise noted under the sample picture.

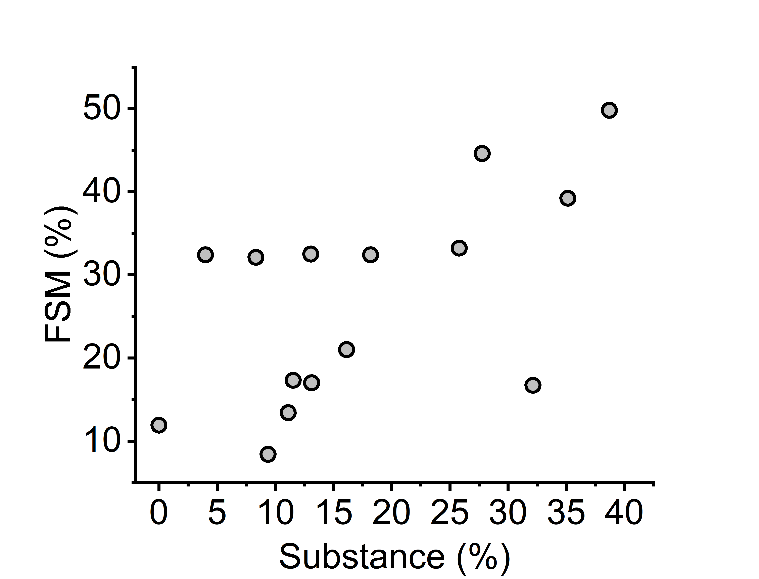

**Figure S2**. The percentage of illicit substances identified from samples *vs* the reported percentage of free school meals. Note we only include schools where there were > 20 samples submitted.

Table S1. Summary of sampling efforts in the academic year 2024/2025

|  | Total samples | SC | THC | Other drug |
| --- | --- | --- | --- | --- |
| Essex 1 (Jan 2025) | 174 | 5 – 2.9% | 1 – 0.6% | 0 |
| Essex 2 (June 2025) | 572 | 14 – 2.4% | 4 – 0.7% | 0 |
| D+C1 (October 2024) | 51 | 4 – 7.8% | 0 – 0% | 2 (ketamine and MDMA) – 4.0% |
| D+C2 (June 2025) | 205 | 10 – 4.9% | 2 – 1.0% | 0 |
| Merseyside (December 2024) | 207 | 2 – 1.0 % | 1 – 0.5% | 0 |
| Lancashire (July 2025) | 215 | 58 – 27.0% | 4 – 1.9% | 1 (ketamine) – 0.5% |

### Table S2. Instagram data.

| Description | SC | THC | unclear | UK ref / postal | Potency | Authentic | logistics | private channel | Trust/Legit | Price/Value | OpSec |
| --- | --- | --- | --- | --- | --- | --- | --- | --- | --- | --- | --- |
| "thc vape liquid, uk plug, verified, #thc nothing on my page is for sale wink emoji, 9:1, Dm for telegram" | 1 |  |  | 1 | x |  |  | x | x |  | x |
| "Flavoured Liquids______, lergit, dilute vape liquids, 10mls and 20ml and more, bulk" | 1 |  |  |  |  | x |  |  | x | x |  |
| bulk deals available DM me to discuss / postage is available but collection is preferred if possible. | 1 |  |  | 1 |  |  | x | x |  | x |  |
| "THC liquid __, we do ship worldwide, tap the link below for your orders, telegram link" | 1 |  |  | 1 |  |  | x | x |  |  |  |
| XXXXX | 1 |  |  |  |  |  |  |  |  |  |  |
| "XXXX, old account got deleted, liquid gold, nothing for sale, IE, next day, follow the link join, telegram link." | 1 |  |  | 1 |  | x | x | x |  |  | x |
| "New account, uk based, nothing for sale, telegram link (XXXX)" | 1 |  |  | 1 |  |  |  | x |  |  | x |
| "THC liquid lokal & import, back up account @venemous.ID, premium e-liquid synthetic import and handmade, daily information #XXXX, dm for bussiness inquiries" | 1 |  |  | 1 |  | x |  | x |  |  |  |
| "XXXXX, GB UK, we are located around the UK, this page is purley for thc oil only and edibles, pm for further information, postage only! No refunds" | 1 |  |  | 1 |  |  | x | x |  |  |  |
| best quality mix and strongest raw in south | 1 |  |  |  | x |  |  |  |  |  |  |
| "LONDON THC VAPE SHOP, 100% THC vape oil straight from the bud, made in the uk GB / US US, delivery/ship, top quality strains from good breeders" | 1 |  |  | 1 |  | x | x |  |  |  |  |
| "XXXXX, next day posting nothing for sale" | 1 |  |  | 1 |  |  | x |  |  |  | x |
| "Magic vape juice UK XXX, £5 postage, nationwide shipping, all parcels out 3PM, DM for any cart or juice related enquiry, synthetic" | 1 |  |  | 1 |  | x | x | x |  | x |  |
| "XXX, thc isolate - made and produced in the UK, varying flavours, can dilute up to 70/30 with normal juice and still feel effects. Contact for details" | 1 |  |  | 1 | x | x |  | x |  |  |  |
| thc liquid Uk | 1 |  |  | 1 |  |  |  |  |  |  |  |
| "THC vape liquid Uk ____, E-Cigarette Store, Tracked 24 hour delivery __ , Strongest liquids on the market , Raw and mixed bottles available, DM FOR PRICE LIST, XXXX.com" | 1 |  |  | 1 | x |  | x | x |  | x |  |
| "THC JUICE__, 24/7 based, Best flavours in town, Shoot us a dm for menu __" | 1 |  |  | 1 |  | x |  | x |  |  |  |
| "THC LIQUID, best stuff going __ collection/post available pics available date time whatever come get ur stuff" | 1 |  |  | 1 |  |  | x |  | x |  |  |
| THC VAPE LIQUID | 1 |  |  |  |  |  |  |  |  |  |  |
| "XXXX, drop off all around bradford, £20 raw" | 1 |  |  | 1 | x |  | x |  |  | x |  |
| Snap- XXXX__ | 1 |  |  |  |  |  |  | x |  |  |  |
| "Disclaimer This chat DOES NOT promote or encourage any illegal activities, all information and contents provided by this channel is meat for education" | 1 |  |  |  |  |  |  |  |  |  | x |
| "XXXX, BEST THC IN UK POSTAL AVAILABLE ANYWHERE__" | 1 |  |  | 1 |  |  | x |  |  |  |  |
| "Your THC vape juice plug, Thc infused vape liquid works with any vape! Message me for details flavours and prices" | 1 |  |  |  |  | x |  | x |  | x |  |
| "XXX, THC liquid" | 1 |  |  |  |  |  |  |  |  |  |  |
| "XXXX, Product/service,Highest Grade THC Vape Liquids,Collections/Delivery,Next Day Delivery Postals,DM me for enquiries... " | 1 |  |  | 1 | x |  | x | x |  |  |  |
| T:H:C vape liquid | 1 |  |  | 1 |  |  |  |  |  |  |  |
| "CBD Liquid, Potent CBD, no traces on drug tests, banging flavours!, DM for more info!, DM for more info!, £35, Based in EastHam and Dagenham" | 1 |  |  | 1 | x | x |  | x |  | x |  |
| "NRW D†SSELDORF____, E-liquid zum entspannen__, legaler Wirkstoff. Nicht nachweisbar.! 2021" | 1 |  |  |  |  |  |  |  |  |  |  |
| "The Liquid Man__, Best Liquid In Townª_, 10mls To Litres , Maximum Potency, Drop Offs On Bulk ___, Dm For Enquiries ____" | 1 |  |  |  | x |  | x | x |  | x |  |
| Drops off anywhere | 1 |  |  |  |  |  | x |  |  |  |  |
|  | 1 |  |  | 1 |  |  |  |  |  | x |  |
| "XXXX, __ 10/10 EST 2017, __ DM to purchase, __ Active on deliveries all over Newham, Tower Hamlets, Barking & Dagenham, Redbridge, Havering & more" | 1 |  |  | 1 |  |  | x | x |  |  |  |
| "XXXX, E-Liquid THC es la manera mas discreta y sana deÊconsumirÊcannabis ya que es un producto fuerte y puro, para los consumidores con fines recreativos" | 1 |  |  |  | x |  |  |  |  |  |  |
| "XXXX, Nothing for sale __, Join the telegram __, | 1 |  |  |  |  |  |  | x |  |  | x |
| Snapchat - XXXX | 1 |  |  | 1 |  |  |  | x |  |  |  |
| "THC_E-liquids them/theirs, Nothing for sale __, DM for WhatsApp/Snap __, UK Deliveries ____" | 1 |  |  | 1 |  |  | x | x |  |  | x |
| "Thc E-Liquid ______, PostalÕs __ - DM for more :" | 1 |  |  | 1 |  |  | x | x |  |  |  |
| "XL litres, UK - NOTHING FOR SALE __ ____ *+44XXXX2* ALL ORDERS HERE telegram @XXX, XXXX | 1 |  |  | 1 |  |  | x | x |  | x | x |
| "Rocket juices__, Check our page out!/UK Based/Nothing for sale/home cooked ____/ delivery available worldwide!" | 1 |  |  | 1 |  | x | x |  |  |  | x |
| "THC e-Liquid Life So ____, __THC Fla__ours __ ______ NoSales, SC: XXXX | 1 |  |  |  |  | x |  | x |  |  | x |
| "ONLY PLACE FOR YOUR LIQUIDS, Fire flavours for THC #1 top flavour STRAWBERRY GRAPE __ __ #2 BLUE SLUSHIE - DM for orders and enquires" | 1 |  |  |  |  | x |  | x |  |  |  |
| Telegram link | 1 |  |  |  |  | x |  | x |  |  |  |
| THC E - LIQUID💧💯 DM for more info📲 - postals available📦📥 | 1 |  |  | 1 |  |  |  | x |  |  |  |
|  | 1 |  |  |  |  |  |  |  |  |  |  |
| "UK based GB, accepting cash app and bank transfer only, telegram link" |  | 1 |  | 1 |  |  | x | x |  |  |  |
| "THC VAPE JUICE, THC empire, your plugs plug, free samples, telegram link" |  | 1 |  |  |  |  |  | x |  | x |  |
| "THC VAPE PENS UK, follow for dm of telegram link" |  | 1 |  | 1 |  | x |  | x |  |  |  |
| Vape thc only in uk |  | 1 |  | 1 |  |  |  |  |  |  |  |
| "THC VAPE PENS, cartidges, uk usa, ireland, australia new zealand" |  | 1 |  | 1 |  | x |  |  |  |  |  |
| "Liquid Gold, Official page for Liquid Gold Thc Cartridges, OMMA Licensed __, Oklahoma City, OK, (Nothing for sale)" |  | 1 |  |  |  | x |  |  | x |  | x |
| "Thc Liquid, snap @XXXX, over 18s only, worldwide postals, Dm for more information, telegram link" |  |  | 1 | 1 |  |  | x | x |  |  | x |
| "THC LIQUID__, worldwide shipping" |  |  | 1 | 1 |  |  | x |  |  |  |  |
| "thc oil, us cali buds, cali distilate edibles, telegram link (XXXX)" |  |  | 1 | 1 |  | x |  | x |  |  |  |
| "medical lab, telegram link" |  |  | 1 |  |  |  |  | x |  |  |  |
| "Strongest Around_____, strongest ""cbd"" vape liquid around, nothing for sale, uk postals, the best flavours, dm me for mor info, telegram link (XXXX)" |  |  | 1 | 1 | x | x | x | x |  |  | x |
| THC-liquid |  |  | 1 |  |  |  |  |  |  |  |  |
| "Thc Liquids, strongest and tastiest bottles on the market, press link below and find out for yourself, telegram link." |  |  | 1 |  | x | x |  | x |  |  |  |
| "THC XXXX LIQUID ____, delivery around the world" |  |  | 1 | 1 |  |  | x |  |  |  |  |
| "SLOW FEET DONT EAT, postals worldwide, lab tested, delta 9, disposables" |  |  | 1 | 1 |  | x | x |  | x |  |  |
| thc Liquid Bottles |  |  | 1 |  |  |  |  |  |  |  |  |
| "German language, 10ml 10 euro" |  |  | 1 |  |  |  |  |  |  | x |  |
| "XXXX, welcom to my cahnnel, TD legit, here you will find everything you need to enjoy, clik to join my channel, website link" |  |  | 1 | 1 |  | x |  | x | x |  |  |
| THC Vape UK.USA |  |  | 1 | 1 |  |  |  |  |  |  |  |
| "US, real stregth real flavour real fast, thc liquid and cali pens that slap, next day uk delivery, telegram link" |  |  | 1 | 1 | x | x | x | x |  |  |  |
| XXXX |  |  | 1 | 1 |  |  |  |  |  |  |  |
|  |  |  | 1 | 1 |  | x |  |  |  |  |  |
| "XXXX, Deliveries & shipping available! 24/7" |  |  | 1 | 1 |  | x | x |  |  |  |  |
|  |  |  | 1 | 1 |  |  |  |  |  |  |  |
| THC-liquid |  |  | 1 |  |  |  |  |  |  |  |  |
|  |  |  | 1 |  |  |  |  |  |  |  |  |
| THC LIQUID BORO |  |  | 1 |  |  |  |  |  |  |  |  |
| "old account got closed nothing for sale __, add snap for info" |  |  | 1 | 1 |  |  |  | x |  |  | x |
|  |  |  | 1 |  |  |  |  |  |  |  |  |
| XXXX |  |  | 1 |  |  |  |  |  |  |  |  |
|  |  |  | 1 |  |  |  |  |  |  |  |  |
| XXXX |  |  | 1 |  |  |  |  |  |  |  |  |
|  |  |  | 1 | 1 |  |  |  |  |  |  |  |
|  |  |  | 1 |  |  |  |  |  | x |  |  |
|  |  |  | 1 |  |  |  |  |  |  |  |  |
| "biscotti thc oil Even, Biscotti thc oil, West belfast, 30 a 10 ml, ______" |  |  | 1 | 1 |  | x |  |  |  | x |  |
| vape thc liquid backup |  |  | 1 |  |  |  |  |  |  |  |  |
|  |  |  | 1 |  |  |  |  |  |  |  |  |
| Total | 45 | 6 | 32 | 48 | 12 | 25 | 27 | 37 | 7 | 13 | 14 |
| % | 0.54 | 0.07 | 0.39 | 0.58 | 0.14 | 0.30 | 0.33 | 0.45 | 0.08 | 0.16 | 0.17 |

**Table S3**. TikTok data.

| Description | SC | THC | unclear | UK ref / postal | Potency | Authentic | logistics | private channel | Trust/Legit | Price/Value | OpSec |
| --- | --- | --- | --- | --- | --- | --- | --- | --- | --- | --- | --- |
| KO RAW THC 1:9 BEAT ANY PLUGS PRICE AND STRENGTH GUARANTEED!!__ nothing4sale__ | 1 |  |  |  | x |  |  |  |  | x | x |
| Telegram link | 1 |  |  | 1 |  |  |  | x |  |  |  |
| THC LIQUID seller ring the bell WhatsApp +44XXXX Snapchat@XXXX | 1 |  |  | 1 |  |  |  | x |  |  |  |
| To Order Contact Me On______________ Telegram __ @XXXX | 1 |  |  | 1 |  |  |  | x |  |  |  |
| Nothing For Sale Here DM for Telegram Link //// | 1 |  |  |  |  |  |  | x |  |  | x |
| We got power they ainÕt got access to. FREE SAMPLES__ @XXXX on Signal | 1 |  |  | 1 | x |  |  | x |  | x |  |
| ____ https://XXXXshop Telegram: XXXX ____ delivery. | 1 |  |  | 1 |  |  | x | x |  |  |  |
| "Message me to buy 10ml-100ml, germany UK" | 1 |  |  | 1 |  |  | x |  |  |  |  |
| Telegram; @XXXX Discord; @XXXX More business on telegram _ | 1 |  |  | 1 |  |  |  | x |  |  |  |
| Click link on my bio to join my telegram channel to order | 1 |  |  |  |  | x |  | x |  |  |  |
| Telegram: @XXXX Discord: @XXXX More details on telegram ____ | 1 |  |  | 1 |  |  |  | x |  |  |  |
| SC **: XXXX | 1 |  |  |  |  |  |  | x |  |  |  |
| Telegram link | 1 |  |  |  |  |  |  | x |  |  |  |
| OLD ACCOUNT GOT LOCKED OFF _ BACK BETTER THAN EVER TELEGRAM: @XXXX | 1 |  |  | 1 |  |  |  | x | x |  |  |
| NOTHING HERE ON TT IS FOR SALE __ BUSINESS ON TELEGRAM Telegram: @XXXX | 1 |  |  |  |  | x |  | x |  |  | x |
| Finest litres around _ Special delivery Telegram link | 1 |  |  | 1 |  |  | x | x |  | x |  |
| Snapchat: XXXX | 1 |  |  |  |  |  |  | x |  |  |  |
| Telegram- @XXXX //XXXX 10ml to Liters to Gallons __ postage | 1 |  |  | 1 |  |  | x | x |  | x |  |
|  | 1 |  |  | 1 |  |  |  |  |  |  |  |
|  | 1 |  |  | 1 |  |  |  |  |  |  |  |
| UK TO UK POSTAGE VERIFIED UK POSTAGE ROYAL MAIL 24H TRACKED | 1 |  |  | 1 |  |  | x |  | x |  |  |
|  | 1 |  |  | 1 |  |  |  |  |  |  |  |
| Based in Bradford Your__ choose your strength__ | 1 |  |  | 1 | x |  |  |  |  |  |  |
| SHIPPING ALL OVER ____ DM FOR TELE __ Private message for XXXX sample. | 1 |  |  | 1 |  |  | x | x |  | x |  |
|  | 1 |  |  | 1 |  |  |  |  |  |  |  |
| UK TO UK POSTAGE VERIFIED UK POSTAGE ROYAL MAIL 24H TRACKED | 1 |  |  | 1 |  |  | x |  | x |  |  |
| BEST SHOP IN THE GAME POWER RAW 11/10 MIXED BOTTLES SHIPPING AVAILABLE ANYWERE | 1 |  |  | 1 | x |  | x |  |  |  |  |
| Hmu people Postals only __ | 1 |  |  | 1 |  |  | x |  |  |  |  |
| UK TO UK POSTAGE VERIFIED UK POSTAGE ROYAL MAIL 24H TRACKED | 1 |  |  | 1 |  |  | x |  | x |  |  |
| Telegram link | 1 |  |  |  |  |  |  | x |  |  |  |
| "THC XXXX, Telegram- XXXX Snapchat**- XXXX" | 1 |  |  | 1 |  |  |  | x |  |  |  |
| "No bio yetTAP IN FOR ONE OF THE BEST PROMOS, BECOMEA KING WITH XXXX KING!" | 1 |  |  | 1 | x |  | x |  |  | x |  |
| raw 10ml £30 | 1 |  |  |  | x |  |  |  |  | x |  |
| SNAP:XXXX _______ STRONGEST BOTTLES 3 puff ko Entertainment purposes only | 1 |  |  | 1 | x |  |  | x |  |  | x |
|  | 1 |  |  |  |  |  |  |  |  |  |  |
| Snapchat XXXX Telegram XXXX Join us now | 1 |  |  | 1 |  |  |  | x |  |  |  |
|  | 1 |  |  | 1 |  |  |  |  |  |  |  |
| ADD THE SNAP XXXX ADD THE TELEGRAM XXXX CEO CHAMP ______ | 1 |  |  | 1 | x |  |  | x |  |  |  |
|  | 1 |  |  | 1 |  |  |  |  |  |  |  |
| Instagram: XXXX ______ | 1 |  |  | 1 |  |  |  | x |  |  |  |
| BEST RAW IN THE UK*_ 10ML-100L COME SHOP WITH THE SOCKET __ NOTHING IS FOR FREE | 1 |  |  | 1 | x |  |  |  |  | x | x |
| Telegram link | 1 |  |  | 1 |  |  |  | x |  |  |  |
| Postals All Over__ DM For Samples IG-XXXX Signal-XXXX__ | 1 |  |  | 1 |  |  | x | x |  | x |  |
|  | 1 |  |  | 1 |  |  |  |  |  |  |  |
|  | 1 |  |  |  |  |  |  |  |  |  |  |
| Insta - XXXXSnap - XXXX FIRST TIME BUYERS £10 A RAW | 1 |  |  |  | x |  |  | x |  | x |  |
| UK safe ____ Guaranteed services give me a follow and I will forward my website | 1 |  |  | 1 |  | x |  | x | x |  |  |
| UK TO UK POSTAGE VERIFIED UK POSTAGE ROYAL MAIL 24H TRACKED TELE@XXXX | 1 |  |  | 1 |  |  | x | x | x |  |  |
| "No goofy moves I keep it loyal with any loyal hommie whoÕs tryna make it, Telegram link " | 1 |  |  |  |  |  |  | x | x |  |  |
| UK TO UK POSTAGE AVAILABLE UK POSTAGE ROYAL MAIL 24H TRACKED GET YOUR ORDERS IN | 1 |  |  | 1 |  |  | x |  |  |  |  |
|  | 1 |  |  | 1 |  |  |  |  |  |  |  |
| Pure thc dm me if your serious no time wasters Best about __ Brum__£30 10mL | 1 |  |  | 1 | x |  |  |  | x | x |  |
|  | 1 |  |  |  |  |  |  |  |  |  |  |
| Telegram DM..__ @XXXX | 1 |  |  | 1 |  |  |  | x |  |  |  |
|  | 1 |  |  |  | x |  |  |  |  |  |  |
| Versand oder Abholung alles mšglich das StŠrste liq im Land +49 XXXX | 1 |  |  |  | x |  | x |  |  |  |  |
| Telegram XXXX | 1 |  |  |  |  |  |  | x |  |  |  |
| NOTHING HERE ON TT IS FOR SALE BUSINESS ON TELEGRAM Telegram: @XXXX | 1 |  |  | 1 |  |  |  | x |  |  | x |
| NOTHING HERE ON TIKTOK IS FOR SALE BUSINESS TELEGRAM ON TELEGRAM @XXXX | 1 |  |  | 1 |  |  |  | x |  |  | x |
| Add via telegram XXXX | 1 |  |  | 1 |  |  |  | x |  |  |  |
| CLICK LINK ON TELEGRAM TO ALL ME UP ON TELEGRAM MAKE SURE YOU ADD UP TELEGRAM | 1 |  |  | 1 |  |  |  | x |  |  |  |
| UK TO UK POSTAGE VERIFIED UK POSTAGE ROYAL MAIL 24H TRACKED TELE@XXXX | 1 |  |  | 1 |  |  | x | x | x |  |  |
| NOTHING HERE ON TT IS FOR SALE BUSINESS ON TELEGRAM/WTSAPP TELE: @XXXX | 1 |  |  | 1 |  |  |  | x |  |  | x |
| hey guys join us through our telegram channel link below for more menu ________ | 1 |  |  | 1 |  | x |  | x |  |  |  |
| click link____on my bi | 1 |  |  | 1 |  |  |  | x |  |  |  |
| UK TO UK POSTAGE VERIFIED UK POSTAGE ROYAL MAIL 24H TRACKED | 1 |  |  | 1 |  |  | x |  | x |  |  |
| "WhatsApp +44XXXX to place order UKÕs #1 STORE __ UK2WW POSTALS __ _____, telegram link" | 1 |  |  | 1 |  |  | x | x |  |  |  |
| Tele - TTHIGH1 Hit me up if you need anything __ Single Or Bulk I Got You __ | 1 |  |  |  |  |  |  | x |  | x |  |
| TELEGRAM: @XXXX | 1 |  |  |  |  |  |  | x |  |  |  |
| Nothing for sale £ All for entertainment purposes __ Message for more informati | 1 |  |  | 1 |  |  |  | x |  |  | x |
| massage me on Telegram @XXXX________ | 1 |  |  | 1 |  |  |  | x |  |  |  |
| Add up via snap ____ Add up via telegram __ | 1 |  |  | 1 |  |  |  | x |  |  |  |
| BEST RAW IN THE UK🧪 10ML-100L COME SHOP WITH THE SOCKET 🔌 NOTHING IS FOR FREE | 1 |  |  | 1 | x |  |  |  |  |  |  |
|  | 1 |  |  | 1 |  |  |  |  |  |  |  |
| DELIVERY THROUGH ROYAL MAIL DISCRET NEXTDAY TELEGRAM @XXXX | 1 |  |  | 1 |  |  | x | x | x |  |  |
| NOTHING ON TIKTOK IS FOR SALE MASSAGE ME ON TELEGRAM TELEGRAM: @XXXX | 1 |  |  | 1 |  |  |  | x |  |  |  |
| LIVERPOOL BASE 🇬🇧🚚🥇✅ MESSAGE ME NOW DIRECTLY TO SHOP ⬇️⬇️⬇️ XXXX, chat.whatsapp.com/XXXX | 1 |  |  | 1 |  |  |  | x |  |  |  |
| signal.group/#XXXX | 1 |  |  | 1 |  |  |  | x |  |  |  |
| old account banned 🚫 For anything send me a dm👨‍🍳 🇬🇧 telegram:👇 @XXXX | 1 |  |  | 1 |  |  |  | x |  |  |  |
| Snap ~ XXXX Tele ~ XXXX | 1 |  |  | 1 |  |  |  | x |  |  |  |
| UK CLONIN🇬🇧💨 ALWAYS ACTIVE PLACE YOUR ORDERS AND YOU HAVE YOU ASAP DELIVERY IN THE UK 🇬🇧✈🚐 | 1 |  |  | 1 |  |  | x |  | x |  |  |
| CLICK LINK BELOW TO JOIN TELEGRAM CHANNEL FOR SHOPPING |  | 1 |  |  |  |  |  | x |  |  |  |
| nothing for sale here join my telegram channel and shop |  | 1 |  |  |  |  |  | x |  |  | x |
|  |  | 1 |  |  |  |  |  |  |  |  |  |
|  |  | 1 |  |  |  |  |  |  |  |  |  |
|  |  | 1 |  |  |  |  |  |  |  |  |  |
|  |  | 1 |  |  |  |  |  |  |  |  |  |
|  |  | 1 |  | 1 |  | x |  |  |  |  |  |
| "King THC dealer, Snapchat-XXXX Telegram-XXXX" |  | 1 |  | 1 |  |  |  | x |  |  |  |
|  |  | 1 |  | 1 |  |  |  |  |  |  |  |
| Disclaimer everything in this group is for art purposes only Join my telegram__ |  | 1 |  | 1 |  |  |  | x |  |  | x |
| Telegram link |  | 1 |  |  |  |  |  | x |  |  |  |
|  |  | 1 |  |  | x |  |  |  |  |  |  |
| Telegram link |  | 1 |  | 1 |  |  |  | x |  |  |  |
| Telegram link |  | 1 |  | 1 |  |  |  | x |  |  |  |
|  |  | 1 |  | 1 |  |  |  |  |  |  |  |
|  |  | 1 |  | 1 |  |  |  |  |  |  |  |
|  |  | 1 |  | 1 |  |  |  |  |  |  |  |
| Telegram link |  | 1 |  | 1 |  |  |  | x |  |  |  |
|  |  | 1 |  | 1 |  |  |  |  |  |  |  |
| Telegram link |  | 1 |  | 1 |  |  |  | x |  |  |  |
| Sc: XXXX Collection__Delivery** |  |  | 1 | 1 | x |  | x | x |  |  |  |
|  |  |  | 1 |  |  |  |  |  |  |  |  |
| AVAILABLE PUPPIES ___ |  |  | 1 | 1 |  |  |  |  |  |  |  |
| UK TO UK POSTAGE VERIFIED UK POSTAGE ROYAL MAIL 24H TRACKED |  |  | 1 | 1 |  |  | x |  | x |  |  |
| "SHIPPING ALL OVER ____ DM FOR TELE AND SNAP__" |  |  | 1 | 1 |  |  | x | x |  |  |  |
|  |  |  | 1 | 1 |  |  |  |  |  |  |  |
| "Flavours on Flavours __ Free postage __ Telegram @uksthc __ The one and only , Telegram link |  |  | 1 | 1 |  | x | x | x | x | x |  |
| TELEGRAM TO ORDER ____ DM @XXXX __________ |  |  | 1 | 1 |  |  |  | x |  |  |  |
| UK THC VAPELIQUID_____ |  |  | 1 | 1 |  |  |  |  |  |  |  |
| NEW ACCOUNT__JOIN TELEGRAM_______ACTIVE AGAIN Telegram link |  |  | 1 | 1 |  |  |  | x | x |  |  |
| Telegram link |  |  | 1 | 1 |  |  |  | x |  |  |  |
| "LIVERPOOL BASE, DELIVERY NEXT DIRECTLY MESSAGE ON TELEGRAM ____ @XXXX" |  |  | 1 | 1 |  |  | x | x |  |  |  |
| click the link ______ to join our telegram shop |  |  | 1 |  |  |  |  | x |  |  |  |
| Telegram main shop __________ Telegram link |  |  | 1 | 1 |  |  |  | x |  |  |  |
|  |  |  | 1 | 1 |  |  |  |  |  |  |  |
|  |  |  | 1 | 1 |  |  |  |  |  |  |  |
|  |  |  | 1 | 1 |  |  |  |  |  |  |  |
| NO SALES❌ CLICK LINK BELOW TO JOIN MY TELEGRAM 👇👇👇👇👇 t.me/XXXX |  |  | 1 | 1 |  |  |  | x |  |  | x |
| City With Gas⛽🇬🇧🇨🇮🏴󠁧󠁢󠁥󠁮󠁧󠁿🏴󠁧󠁢󠁳󠁣󠁴󠁿 Telegram @XXXX |  |  | 1 | 1 | x |  |  | x |  |  |  |
| Totals | 81 | 20 | 19 | 91 | 17 | 6 | 25 | 71 | 15 | 13 | 12 |
| % | 0.68 | 0.17 | 0.16 | 0.76 | 0.14 | 0.05 | 0.21 | 0.59 | 0.13 | 0.11 | 0.10 |

Table S4. Thematic analysis coding framework

| Theme | Description | Example Codes | Illustrative Quote |
| --- | --- | --- | --- |
| Extreme Potency | Language that emphasizes an overwhelming, powerful, or incapacitating effect of the product, often as the primary selling point. | KO, Strongest, Raw, Beast, Madness, Punch, 1:9 | "KO RAW THC 1:9 BEAT ANY PLUGS PRICE AND STRENGTH GUARANTEED!!🔥" |
| Product Form/Authenticity | Language that describes the physical characteristics, flavour, or perceived quality of the product, often using terminology from the legitimate cannabis market. | Flavours, Carts, Oil, Resin, Branded, Cali | "Flavours on Flavours 🤤 Free postage 🚚" |
| Logistics & Distribution | Language detailing the methods of delivery, shipping capabilities, and geographic reach of the seller. | Postage, Postals, Shipping, Delivery, Drop off, Tracked, Nationwide, Worldwide | "UK TO UK POSTAGE VERIFIED UK POSTAGE ROYAL MAIL 24H TRACKED" |
| Funneling to Private Channels | Explicit instructions or links directing users to communicate or transact on encrypted, private platforms rather than the public-facing social media page. | Telegram, Snapchat, WhatsApp, DM for tele, link in bio | "CLICK LINK BELOW TO JOIN TELEGRAM CHANNEL FOR SHOPPING" |
| Building Trust & Legitimacy | Language used to reassure potential buyers of the seller's credibility and the reliability of their service, separate from just logistics. | Verified, legit, the one and only, reviews, pics available date time | "UK TO UK POSTAGE VERIFIED UK POSTAGE ROYAL MAIL 24H TRACKED" |
| Price & Value Proposition | Language that focuses on the cost-effectiveness or deals available, positioning the seller as a competitive option in the market. | Beat any plugs price, bulk, deals, offers | "KO RAW THC 1:9 BEAT ANY PLUGS PRICE AND STRENGTH GUARANTEED!!🔥" |
| Plausible Deniability & OpSec | "Wink-and-nod" language used to create a thin veil of deniability about the account's illicit activities and protect it from moderation. | Nothing for sale here, nothing4sale🤫, educational purposes only, over 18s only | "nothing on my page is for sale 😉" |
